# Aerosol Emission During Speech: Investigating the Role of Glottal Configuration and Respiratory Effort

**DOI:** 10.1101/2025.03.16.25324059

**Authors:** Allison Hilger, Tehya Stockman, Corey Murphey, Jacqueline McCurdy, Shelly Miller

**Affiliations:** Department of Speech, Language, and Hearing Science, University of Colorado Boulder, Boulder, Colorado, United States of America; Department of Mechanical Engineering, University of Colorado Boulder, Boulder, Colorado, United States of America; Department of Computer Science, University of Colorado Boulder, Boulder, Colorado, United States of America; Current Employment: Adams County, 4430 S Adams County Pkwy, Brighton, CO 80601; Current Employment: HCA HealthONE Sky Ridge Outpatient Rehabilitation, 10103 RidgeGate Parkway, Suite G01A, Lone Tree, CO 80124

## Abstract

**Introduction:** Speech-driven aerosol generation plays a key role in airborne disease transmission, yet the physiological mechanisms remain poorly understood. While prior research suggests vocal fold vibration contributes to aerosol production, airflow turbulence and glottal configuration may be stronger determinants. This study examines how the type of phonation influences aerosol generation while controlling for respiratory effort.

**Methods:** Five healthy female adults (22–43 years) sustained vowels across six phonation types: modal voicing, glottal fry, falsetto, whispered speech, loud speech, and vowels preceded by /h/. Aerosol concentration and size distribution (0.1–20 µm) were measured using an aerodynamic particle sizer (APS). Laryngoscopy quantified normalized glottal gap, and CO_2_ range was recorded to control for respiratory effort. Bayesian regression models assessed relationships between phonation type, aerosol generation, and physiological predictors.

**Results:** Whispering and loud speech produced the highest aerosol concentrations, while glottal fry generated the least. Smaller aerosol particles (0.1–1 µm) were most prevalent across tasks, highlighting their potential for airborne transmission. Whispering exhibited a bimodal aerosol distribution, with increased emissions at both the smallest (0.1–1 µm) and largest (10–20 µm) particles sizes. Despite the assumption that vocal fold vibration is necessary for aerosol production, whispering, a voiceless phonation, generated the most aerosols, suggesting airflow turbulence and glottal configuration are stronger contributors. Normalized glottal gap was the strongest predictor of aerosol output, followed by CO_2_ range, while harmonics-to-noise ratio had a smaller effect.

**Conclusion:** Vocal fold vibration alone is not necessary for high aerosol generation; turbulent airflow through a partially open glottis is a key driver. These findings have implications for airborne disease transmission, particularly in densely occupied environments. Future research should explore real-world speech patterns to refine strategies for minimizing respiratory particle exposure.

## Introduction

The rapid airborne transmission of SARS-CoV-2 during the COVID-19 pandemic emphasized the importance of understanding how speech and singing contribute to aerosol dispersion, particularly in enclosed spaces such as restaurants, classrooms, and choir settings (1–3). Airborne pathogens such as COVID-19, influenza, and tuberculosis spread through exhaled aerosol particles generated during breathing, coughing, sneezing, and vocalizing (i.e., phonation from speaking or singing) (4–6). Despite the established role of exhaled particles in disease transmission, the specific physiological mechanisms underlying aerosol generation in the respiratory and vocal tracts remain poorly understood. Here we refer to an airborne droplet as a small particle of liquid often containing a virus or bacteria that is released when someone talks or sings. Droplets evaporate rapidly upon emission and the diameter of the droplet shrinks, but this phenomenon is not well understood (7).

When comparing expiratory events, speech and singing generate more particles per unit of time than coughing or breathing, with smaller particles that remain airborne longer and increase the likelihood of inhalation by others (8,9). Louder speech has been shown to generate higher aerosol concentrations than quiet speech, raising the question of whether this effect is driven by increased lung pressure, greater amplitude of vocal fold vibration, or both (8,10).

Interestingly, recent findings indicate that whispering produces more aerosol particles than modal speech, likely due to higher airflow turbulence through a partially open glottis, which increases respiratory particle formation (11,12). This challenges the assumption that quieter vocalization minimizes airborne transmission risk.

Theories of aerosol generation propose that exhaled particles originate from multiple points within the respiratory tract. In the lungs, a fluid film across the bronchioles bursts during exhalation, generating small particles (<1 µm) (9,10,13). In the larynx, particles can form due to vortex shedding and vibration-induced atomization caused by the vibration of the vocal folds (∼0.5-5 µm) (5). In the oral cavity, the opening and closing of the mouth causes the formation of fluid/saliva jets and bubble-bursting, which generate particles that tend to be larger (∼5-20 µm) (9,14–16). These size distinctions are significant, as smaller laryngeal-generated particles (∼0.5-5 µm) remain airborne longer and fit within the size range required for conveying many viral and bacterial particles such as tuberculosis bacilli, COVID-19, and influenza (17–19).

Changes in phonation quality and respiratory effort significantly affect aerosol concentration and emission rates (and possibly droplet size). Loud speech and singing require greater subglottal pressure, initiated by higher lung volumes (∼60-90%VC), which may increase aerosol generation (8,20–22). Voiced sounds (e.g., /b, g, z/) generate more aerosol particles than voiceless sounds (e.g., /p, k, s/), suggesting a role for vocal fold vibration in aerosol production (23). Additionally, breathing patterns alter aerosol emission. Exhaling below functional residual capacity increases aerosol concentration, whereas breath-holding reduces it (10,24). Further, whispering and loud speech generate high aerosol concentrations, reinforcing the importance of airflow dynamics in vocalized aerosol generation (8,11,12,25)

Despite increasing research interest in aerosol generation from speech, critical gaps remain in understanding how phonatory biomechanics and respiratory control interact to influence aerosol output. Many studies have focused on speech intensity and phoneme type, but few have systematically controlled respiratory effort, glottal positioning, or vocal fold vibration periodicity. This study aims to determine which physiological factors most strongly predict aerosol generation during phonation. Specifically, we investigate the relationship between carbon dioxide range (CO_2_ range), glottal gap, and harmonics-to-noise ratio (HNR) to assess their impact on droplet concentration. Each of these measures reflects distinct physiological processes that can influence aerosol generation during vocalization. CO_2_ range serves as an indicator of respiratory effort and lung volume, both of which can affect the velocity and force of exhaled airflow, thereby influencing the number and size of emitted particles. The glottal gap reflects the degree of vocal fold separation (obtained using laryngoscopy), which affects the pressure and airflow dynamics as air passes through the glottis, potentially impacting aerosol formation. HNR, a measure of vocal signal quality, reflects the balance between harmonic and noisy components in vocal output, with higher noise levels suggesting greater turbulence that may contribute to aerosol generation. By comparing these measures, the study aimed to identify the primary physiological mechanisms driving aerosol production during vocalization.

To achieve this, we had study participants produce six distinct phonation conditions (modal voicing, glottal fry, falsetto, whispered speech, loud speech, and the /h/ sound prior to sustained vowel) to evaluate how variations in respiratory effort, glottal positioning, and vocal fold vibration periodicity affect aerosol output. Phonation types vary in their acoustic output and underlying physiological mechanisms, which may influence aerosol generation. Modal voicing was the first phonation quality in which the glottis is fully (or mostly) closed during the glottal cycle, resulting in a clear, periodic vocal waveform (26). The second phonation quality is glottal fry, also known as creaky voice, which is characterized by irregular phonation at the low end of the vocal register (27). Falsetto is the third phonation quality, which perceptually appears high in pitch and is physiologically characterized by thin and stretched vocal folds with more rapid cycles of oscillation (28). The fourth phonation quality is whispered speech in which the glottis is partially open creating turbulent airflow through the smaller opening, and is perceptually realized as a breathy, aperiodic phonation quality (29). Fifth is loud speech in which a clear voice is used but at 10 dB SPL louder than comfortable voicing in this study. Finally, the last phonation quality is saying /h/ prior to each vowel, for example, “heeeee” or “haaaaa.” This final voicing style is produced with a partially open glottis for the /h/ sound followed by the fully closed glottis for the sustained vowel. These six phonation qualities were chosen to measure how changes in glottal positioning, vocal fold oscillation periodicity, and airflow influence particle concentration and size. For example, the glottis is fully closed for modal voicing, partially closed for glottal fry, and partially open for whispered speech and the /h/ preceding the vowel. Oscillation periodicity is greater for modal voicing, falsetto, and loud voicing, and reduced for glottal fry, whispered speech, and the /h/ preceding the vowel. Finally, airflow is predicted to be greatest for loud speech based on prior work that greater lung volume is used to speak louder (30). We then predict that airflow will be greater for whispered speech, followed by modal voicing, falsetto, /h/ preceding the vowel, and (the least amount) glottal fry. Overall, this investigation is essential for understanding the factors that contribute to airborne particle emissions, particularly in contexts where vocal communication may increase the risk of respiratory disease transmission.

## Methods

### Participants

This experiment included five healthy female participants, all between the ages of 22– 43 years (mean = 31), with no history of voice or respiratory impairments. The study was approved by the University of Colorado Boulder Institutional Review Board (IRB Protocol #20-0282) for the aerosol measurements and the Colorado Multiple Institutional Review Board (COMIRB Protocol #22-1678) for the laryngoscopy. Participants were provided with informed written consent. Participant recruitment took place from 01/01/2022 to 01/30/2023.

### Procedures

To measure particle concentration and size across different phonation types, participants produced sustained vowels using six distinct phonation qualities: modal voicing, glottal fry, falsetto, whispered voicing, loud voicing, and vowels preceded by an /h/. Each phonation quality was produced across four sustained vowels, /ɑ/ (“saw”), /i/ (“see”), /ɛ/ (“said”), and /ə/ (“sun”) chosen for their differences in tongue position. The goal was to measure aerosol generation across phonation tasks varying by open vocal tract postures to maximize generalizability. Participants sustained each vowel for five seconds and repeated the production three consecutive times per phonation condition. To ensure consistency, a timed visual presentation guided participants through the sequence of vowel productions and phonation types. Participants vocalized into a funnel connected to an aerodynamic particle sizer (APS, 3321, TSI) to measure particle size distributions for particles that reach the APS sensor at the end of the funnel. The APS measured droplet size distributions of ∼0.1 – 20 µm. Additionally, CO_2_ concentrations were measured using a Licor (LI-7000, Li-Cor, Lincoln, NE), which samples CO_2_ once per second with the data averaged to every minute. The APS measured a size distribution every minute. To reduce background aerosol concentration, two portable HEPA air cleaners (Air Response Air Purifier, Oreck) were used to decrease the background levels of aerosol between runs. The total particle number background concentration in the room was 0.03 - 0.1#/cm3 as reported by the APS.

Participants wore an AKG C420 condenser microphone positioned over the ear, connected to a Motu Ultralite-mk3 Hybrid audio interface. A real-time visual feedback system displayed their speech loudness, helping them maintain a target range of 70–75 dB SPL during phonation. Each sequence of three vowel productions was followed by 35 seconds of nasal breathing, allowing sufficient time for the aerosol plume to dissipate and preventing signal overlap between trials.

In a separate testing session at an otolaryngology clinic, laryngoscopy was used to measure glottal positioning across the six phonation qualities using the normalized glottal area during the open phase of vocal fold oscillation. Nasolaryngoscopy videos were obtained from all five participants while they sustained the /i/ vowel across all six phonation qualities. A speech-language pathologist conducted the laryngoscopy using a flexible laryngoscope (Olympus, Olympus Surgical, Center Valley, PA) inserted transnasally and positioned near the arytenoid cartilages. Another speech-language pathologist captured still images of the open phase of oscillation for three frames per phonation type.

The normalized glottal gap was then measured for each still shot using ImageJ (National Institutes of Health, Bethesda, MD, USA, https://imagej.nih.gov/ij) based on the procedures used in (31). Normalized glottal area was calculated relative to vocal fold length, which was measured from the anterior vocal process to the anterior connection of the vocal folds with the thyroid cartilage. This length was used to set the scale function in ImageJ. The glottal area was then quantified by adjusting image brightness using the color threshold function, isolating the glottic gap. A line sector measurement was used to determine the distance at the widest point of the glottic gap. The resulting normalized glottal gap measure was calculated by dividing the anterior gap by the square of vocal fold length times 100. This process was repeated for three consecutive phonatory cycles, and values were averaged to obtain a final measurement for each participant and phonation type.

### Outcome Measures

The primary outcome measures were droplet concentration and droplet size, measured as dN/dlogDp, obtained from the aerodynamic particle sizer (APS). To ensure a meaningful interpretation of droplet size distributions, droplet concentration was categorized into five size bins: 0.1-1 µm, 1-2.5 µm, 2.5-5 µm, 5-10 µm, and 10-20 µm. These bins were selected based on prior literature that distinguishes between respirable aerosols (<2.5 µm), transition-sized droplets (2.5-10 µm), and larger droplets (>10 µm), which have different aerodynamic behaviors and transmission risks (9,17). Categorizing droplet size enables statistical modeling to evaluate trends across biologically relevant ranges that align with infectious disease transmission models.

To define the analysis window for droplet concentration and size measurements, CO_2_ range was used as an objective criterion. Since CO_2_ levels rise during speech, the gas analyzer provided a method to delineate speech and nasal breathing phases. The minimum and maximum CO_2_ values were calculated for each 75-second trial window (15 seconds of speech, 60 seconds of nasal breathing). The APS analysis window was then set post facto by applying a 25% threshold of this range, ensuring that particle data captured corresponded to active phonation. This analysis window was also used to determine CO_2_ range by subtracting the minimum CO_2_ output from the maximum output. The other measures in this study included normalized glottal gap, described previously, and harmonics-to-noise ratio (HNR). HNR was computed using Praat software (32) from the audio from the laryngoscopy videos to quantify vocal fold periodicity by participant and phonation task.

### Statistical Analysis

The first research question asked how droplet concentration varied by vocal task and droplet size while controlling for CO_2_. Specifically, a Bayesian regression model was implemented using the brms package (33) in R (R version 4.3.2) to analyze the effects of vocal task and droplet size on droplet concentration while controlling for CO_2_ range. The dependent variable was droplet number concentration per diameter size range (dN/dlogDp, #/cm^3^), and the independent variables included vocal task (modal speech, loud speech, whispered speech, glottal fry, falsetto, and /h/ before vowel), particle size (µm), and CO_2_ range. The model included an interaction term between task and size to assess whether differences in droplet concentration between vocal tasks varied across particle sizes. Additionally, CO_2_ range was included as a covariate to account for the potential influence of the amount of air expelled on droplet concentration. CO_2_ range was scaled to improve model convergence and interpretability, ensuring that parameter estimates were on a comparable scale.

A lognormal family was used to account for the right-skewed, positive distribution of droplet concentration, with an identity link function for both the mean (μ) and standard deviation (σ). Weakly informative priors were specified to regularize estimates without overly influencing the results. Specifically, the intercept was assigned a normal prior with a mean of zero and a standard deviation of one, reflecting the assumption that the average particle concentration is centered around zero, with most values expected to fall within a reasonable range. Similarly, all regression coefficients for the predictors were given normal priors with a mean of zero and a standard deviation of one. This weakly informative prior assumes that the predictors are likely to have small to moderate effects on droplet concentration while still allowing for larger effects if supported by the data. Four Markov Chain Monte Carlo (MCMC) chains were run, each with 2000 iterations, including 1000 warm-up iterations, resulting in 4000 post-warm-up draws. The model was fit using the No-U-Turn Sampler (NUTS), an adaptive version of Hamiltonian Monte Carlo (HMC), with a maximum tree depth of 15 and an adaptation delta of 0.9 to improve sampling efficiency and convergence.

To evaluate the influence of the CO_2_ range control variable, a reduced model was constructed by excluding CO_2_ range from the predictors. The performance of the full model, which included CO_2_ range, was compared to the reduced model using leave-one-out cross-validation (LOO). This comparison was conducted using the loo function in the R package brms (33), which calculates the expected log pointwise predictive density (elpd) for each model. The difference in elpd between the two models, along with its standard error, was used to assess whether including CO_2_ range significantly improved the model’s predictive accuracy. The results of this comparison indicated that including CO_2_ range led to a better fit, as evidenced by a higher elpd value for the full model, supporting its inclusion as a control variable in the analysis.

A second Bayesian model was run to determine which mechanism of phonation, CO_2_ range, vocal fold periodicity (HNR), or normalized glottal gap, most robustly predicted particle concentration during vocal tasks while accounting for the lognormal distribution of the data. These predictors were standardized to allow for a relative interpretation of their effects on droplet concentration. Predictors were standardized by the scale() function, resulting in variables with a mean of zero and a standard deviation of one. This standardization facilitates the interpretation of regression coefficients as the expected change in particle concentration for a one-standard-deviation increase in each predictor.

A lognormal family was specified for the response variable, reflecting the right-skewed distribution of droplet concentration data. The lognormal distribution ensures that predicted values remain positive, aligning with the non-negative nature of droplet concentration measurements. The model was fit using the No-U-Turn Sampler (NUTS), a variant of Hamiltonian Monte Carlo, with four Markov chains run in parallel to enhance computational efficiency and ensure convergence. Each chain consisted of 2,000 iterations, with the first 1,000 used as warm-up samples to optimize the sampler.

Weakly informative priors were applied to promote regularization without overly constraining the parameter estimates. Specifically, the intercept was assigned a normal prior with a mean of zero and a standard deviation of one, reflecting the assumption that the baseline droplet concentration is centered around zero. Regression coefficients for the scaled predictors were also assigned normal priors with a mean of zero and a standard deviation of one, indicating that the predictors are expected to have small to moderate effects on droplet concentration.

Model convergence was assessed using the Rhat statistic, with values close to 1.00 indicating good convergence. Effective sample sizes (ESS) were examined to ensure sufficient sampling precision. Posterior distributions of model parameters were summarized using median estimates and 95% highest posterior density (HPD) intervals. Pairwise comparisons between vocal tasks were conducted using the emmeans package with Bonferroni adjustments to control for multiple comparisons. Model fit and predictive accuracy were evaluated using approximate leave-one-out cross-validation (LOO), with Pareto-smoothed importance sampling (PSIS) to assess model diagnostics. For all model results, a robust effect was determined if the HPD interval did not include zero, indicating a statistically meaningful association between the predictor and the outcome variable. Data and code can be found at https://osf.io/uc6km/.

## Results

### Particle Concentration by Task and Size

A Bayesian regression model was used to examine the effects of vocal task and particle size on aerosol concentration while controlling for CO_2_ range, with a lognormal likelihood to account for the skewed distribution of the response variable. The model included task and droplet size as fixed effects, along with their interaction. All Rhat values were equal to 1.00, indicating model convergence and no divergent transitions were detected. The effective sample sizes (ESS) were sufficient for reliable inference, and posterior predictive checks showed that the model accurately captured the observed data distribution.

A Bayesian contrast analysis compared aerosol concentrations across vocal tasks and droplet sizes with 95% HPD intervals. The results confirm that smaller particles (0.1-1 μm) exhibit robustly higher concentrations than all other larger size ranges, including 2.5-5 μm (Estimate = 0.4398, 95% HPD: [0.275, 0.6032]), 5-10 μm (Estimate = 0.5602, 95% HPD: [0.220, 0.8758]), and 10-20 μm, (Estimate = 0.8308, 95% HPD: [0.737, 0.9245]). Among vocal tasks, as seen **in Figure 1**, whispering produced the highest aerosol concentrations, robustly exceeding Glottal Fry (Estimate = 0.4881, 95% HPD: [0.1247, 0.8087]), Falsetto (Estimate = 0.3200, 95% HPD: [0.0516, 0.6124]), and Modal Speech (Estimate = 0.4155, 95% HPD: [0.1734, 0.6744]). Loud speech also produced more particles than modal speech (Estimate = 0.2998, 95% HPD: [0.1367, 0.4939]) and glottal fry (Estimate = 0.3663, 95% HPD: [0.0739, 0.6560]), but was not robustly different from whispering.

**Figure 1:**
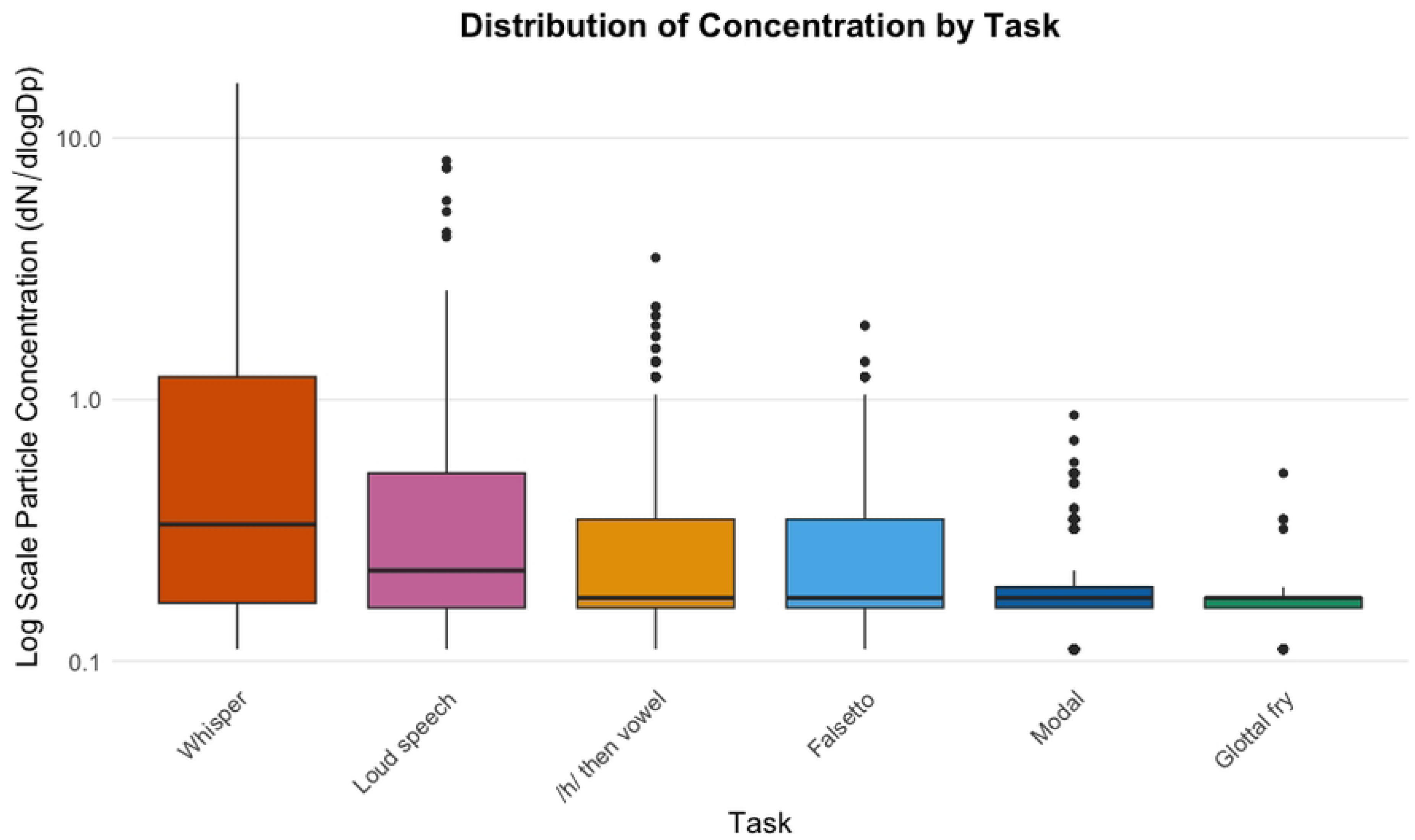
Boxplot showing the distribution of aerosol concentration across different vocal tasks, ordered by mean concentration (lognormal transformation of dN/dlogDp, #/cm^3^). Concentration values are plotted on a log scale to enhance the visualization of differences. Each box represents the interquartile range (IQR), with the horizontal line indicating the median concentration. Whiskers extend to the smallest and largest values within 1.5 times the IQR, while individual points beyond this range represent potential outliers. Colors correspond to distinct vocal tasks for consistency with other analyses

A total of 185 robust interactions were identified in the post hoc pairwise comparisons, where the 95% highest posterior density (HPD) intervals did not include zero. These results indicate robust differences in aerosol concentration across vocal tasks and particle sizes. To highlight the most relevant impactful findings, interaction effects (shown in **Figure 2**), we have summarized the results by task and size as follows.

**Figure 2:**
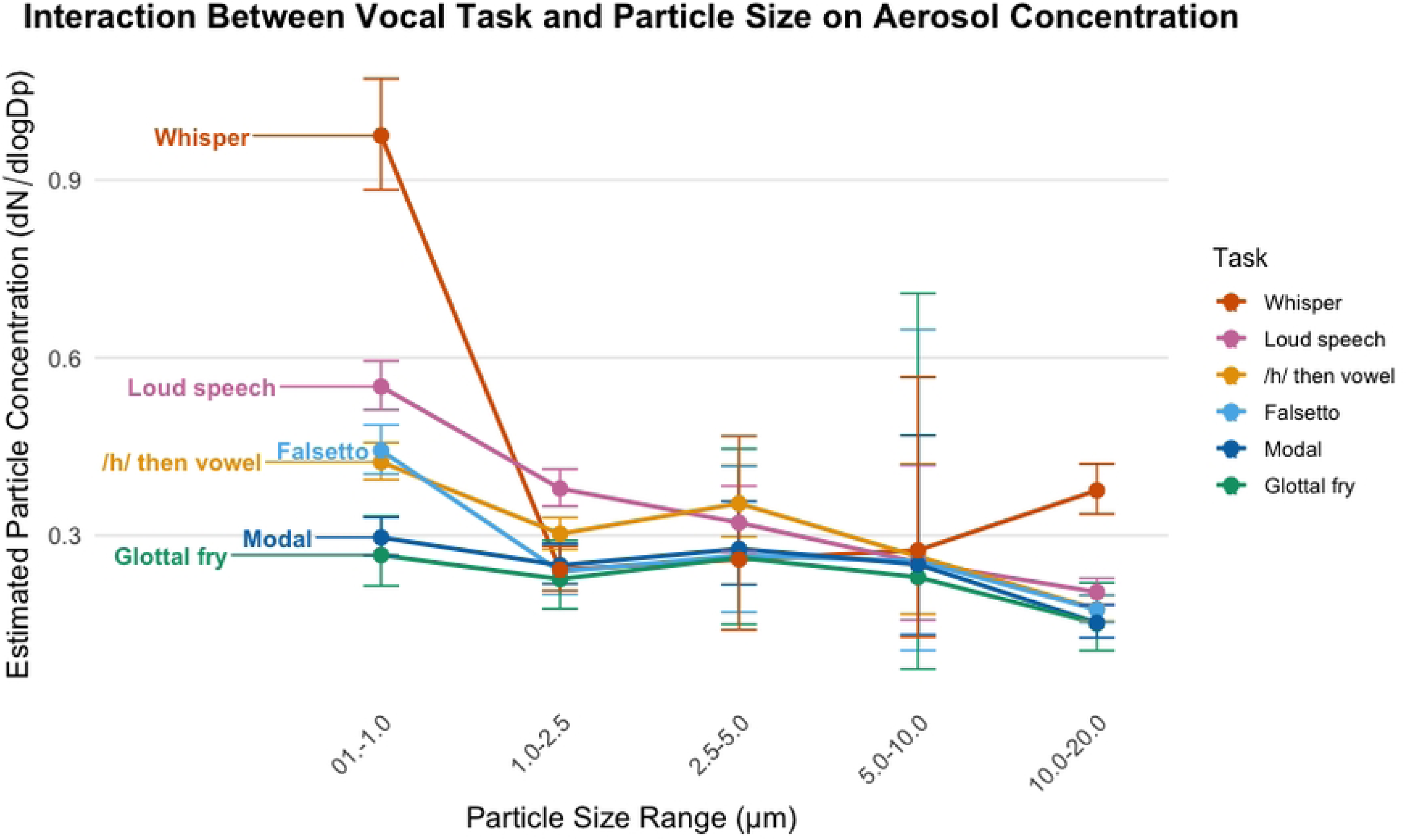
Interaction between vocal task and particle size on aerosol concentration. Estimated aerosol concentration (#/cm^3^) is plotted across five particle size ranges (0.1–1 µm, 1–2.5 µm, 2.5–5 µm, 5–10 µm, 10–20 µm) for each vocal task. Lines connect the estimated means for each task, with error bars representing 95% credible intervals.

### The Interaction of Droplet Concentration Across Particle Size Ranges for Different Vocal Tasks

Pairwise comparisons of the interaction between vocal task and particle size range revealed that concentrations were highest at the smallest particle size range (0.1–1 µm) and progressively declined as droplet size increased for all tasks except whispering. This trend was consistent across loud speech, falsetto, modal speech, /h/ then vowel, and glottal fry, with robust differences between the smallest and largest size ranges.

Whispering exhibited the strongest contrast between size ranges, producing substantially higher droplet concentrations in the 0.1–1 µm range than all other size ranges (βs > 0.94, all 95% HPDs excluding zero). Notably, whispering at 10–20 µm generated more droplets than at 1–2.5 µm (β = 0.4388, 95% HPD = [0.2644, 0.6105]), suggesting a secondary peak at larger particle sizes, which was not observed in other vocal tasks.

For loud speech, particle concentration was robustly higher in the 0.1–1 µm range compared to all larger size ranges, including 1-2.5 µm (β = 0.3763, 95% HPD = [0.2686, 0.4852]), 2.5–5 µm (β = 0.529, 95% HPD = [0.354, 0.720]), 5-10 µm (β = 0.7772, 95% HPD = [0.3254, 1.2920]), and 10–20 µm (β = 0.993, 95% HPD = [0.865, 1.1246]). Additionally, the 1–2.5 µm range generated more particles than the 10–20 µm range (β = 0.6168, 95% HPD = [0.4771, 0.7494]), reinforcing a consistent decrease in particle emissions at larger sizes. Similarly, falsetto showed robustly higher particle concentrations at 0.1–1 µm compared to 1-2.5 µm (β = 0.6154, 95% HPD = [0.4195, 0.8107]), 2.5–5 µm (β = 0.5166, 95% HPD = [0.0616, 0.9816]), and 10–20 µm (β = 0.9313, 95% HPD = [0.7691, 1.0927]), with a significant drop-off at larger particle sizes. The /h/ then vowel task exhibited the same pattern, with the smallest size range producing robustly more droplets than 1-2.5 µm (β = 0.3355, 95% HPD = [0.2288, 0.4041]), 5-10 µm (β = 0.4634, 95% HPD = [0.0408, 0.9515]), and 10–20 µm (β = 0.8853, 95% HPD = [0.746, 1.021]). For modal speech, robust differences were found between the smallest and largest size ranges, with robustly greater particle concentration at 0.1–1 µm than at 1-2.5 µm (β = 0.1699, 95% HPD = [0.0064, 0.3414]) and at 10–20 µm (β = 0.667, 95% HPD = [0.4652, 0.8786]). Glottal fry showed the fewest robust differences but followed the same overall trend, with higher particle concentrations in the 0.1–1 µm range compared to 10–20 µm (β = 0.506, 95% HPD = [0.1289, 0.9885]).

Overall, these findings indicate that most vocal tasks predominantly generate small particles, with the highest concentration observed in the 0.1–1 µm range and progressively fewer emissions at larger droplet sizes. However, whispering uniquely produces substantial emissions across both small and large particle ranges.

### Differences Among Vocal Tasks Within Each Particle Size Range

At the smallest particle size range (0.1–1 µm), whispering consistently produced the highest particle concentration among all tasks, with robust differences compared to every other vocalization. Specifically, whispering generated more particles than /h/ then vowel (β = 0.8315, 95% HPD = [0.7031, 0.9491]), falsetto (β = 0.7846, 95% HPD = [0.6532, 0.9233]), glottal fry (β = 1.2955, 95% HPD = [1.0527, 1.5327]), loud speech (β = 0.5658, 95% HPD = [0.4448, 0.6868]), and modal speech (β = 1.1898, 95% HPD = [1.0417, 1.3294]), with all 95% HPD intervals excluding zero. Beyond whispering, glottal fry produced the lowest particle concentration, robustly lower than /h/ then vowel (β = −0.4648, 95% HPD = [−0.6897, −0.2405]) and falsetto (β = −0.5119, 95% HPD = [−0.7445, −0.269]). Meanwhile, loud speech produced more particles than falsetto (β = 0.218, 95% HPD = [0.1032, 0.3334]) and modal speech (β = 0.6238, 95% HPD = [0.5001, 0.7523]).

At the 1–2.5 µm droplet size range, whispering no longer exhibited robustly higher particle concentrations than other tasks. However, substantial differences emerged between glottal fry and other vocal tasks, with glottal fry producing fewer particles than /h/ then vowel (β = −0.2902, 95% HPD = [−0.5493, −0.0364]) and loud speech (β = −0.5088, 95% HPD = [−0.7701, −0.2676]). Loud speech produced more particles than falsetto (β = 0.487, 95% HPD = [0.299, 0.677]), glottal fry (β = 0.578, 95% HPD = [0.328, 0.848]), modal speech (β = 0.457, 95% HPD = [0.2676, 0.6501]), and whispering (β = 0.4482, 95% HPD = [0.2815, 0.621]).

At the intermediate droplet size ranges (2.5–5 µm and 5–10 µm), differences between vocal tasks were less pronounced, with no robust differences observed as all 95% HPD intervals included zero. However, at the largest particle size range (10–20 µm), whispering again produced robustly more droplets than /h/ then vowel (β = 0.7658, 95% HPD = [0.6033, 0.928), falsetto (β = 0.7665, 95% HPD = [0.6039, 0.9457]), glottal fry (β = 0.9093, 95% HPD = [0.5003, 1.3096]), loud speech (β = 0.6115, 95% HPD = [0.4635, 0.7773]), and modal speech (β = 0.9061, 95% HPD = [0.7036, 1.119]), with all 95% HPD intervals excluding zero. These results indicate that whispering is unique in its high emissions at both the smallest and largest size ranges, while other tasks exhibit a consistent decline in aerosol generation as particle size increases.

### Comparison of Phonation Mechanisms by Droplet Concentration

A Bayesian regression model was used to evaluate the effects of CO_2_ range, HNR, and normalized glottal gap on particle concentration, with all predictors standardized to allow for relative interpretation of their effects (**Figure 3**). The model assumed a lognormal distribution of the response variable, particle concentration, to account for its right-skewed distribution. The intercept was estimated at −1.83 (95% HPD: [−1.87, −1.79]). Among the standardized predictors, the normalized glottal gap had the strongest positive association with particle concentration, with an estimate of 0.51 (95% HPD: [0.47, 0.55]). This result indicates that a one standard deviation increase in the normalized glottal gap is associated with an increase in particle concentration. The standardized CO_2_ range exhibited a positive relationship with particle concentration, with an estimate of 0.27 (95% HPD: [0.23, 0.31]), suggesting that a one standard deviation increase in CO_2_ range is associated with a higher particle concentration. Additionally, HNR showed a modest positive effect on particle concentration, with an estimate of 0.05 (95% HPD: [0.01, 0.10]), indicating that a one standard deviation increase in HNR is associated with a slight increase in particle concentration. Overall, the model indicates that the standardized normalized glottal gap and CO_2_ range are robust predictors of particle concentration, with the CO_2_ range reflecting respiratory drive and glottal gap capturing phonatory efficiency, with HNR playing a smaller but still meaningful role, reflecting vocal fold periodicity. **Figure 4** reflects differences in these variables by task.

**Figure 3:**
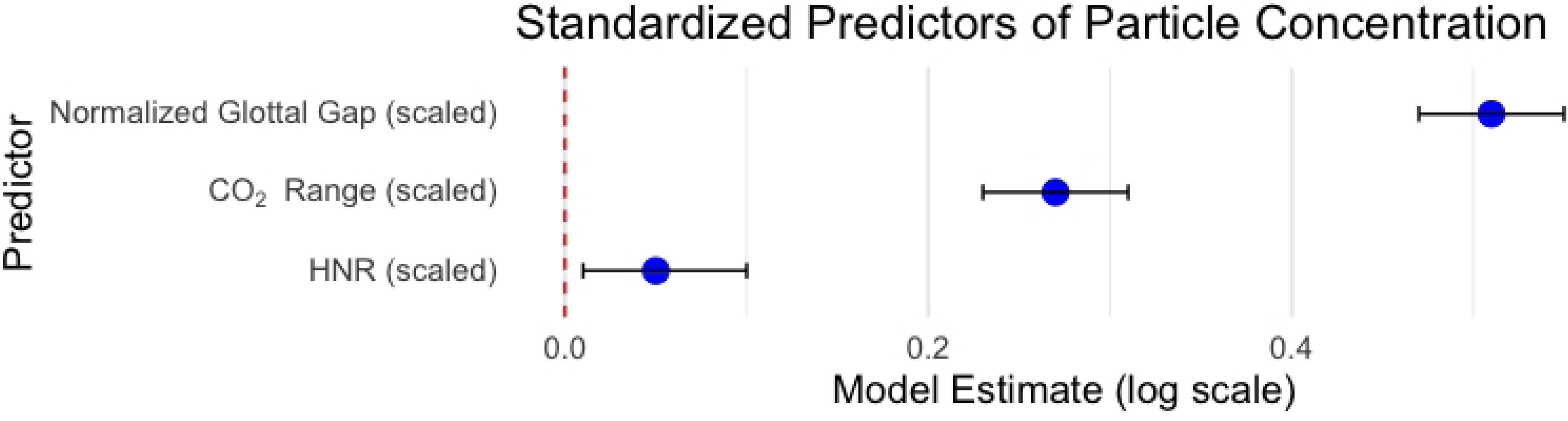
Forest plot illustrating the standardized regression coefficients for predictors of droplet concentration. Point estimates are shown as blue dots, with black horizontal lines representing 95% highest posterior density intervals. The red dashed vertical line at zero indicates no effect. Predictors include CO_2_ Range (scaled), HNR (scaled), and Normalized Glottal Gap (scaled), all standardized for relative interpretation.

**Figure 4:**
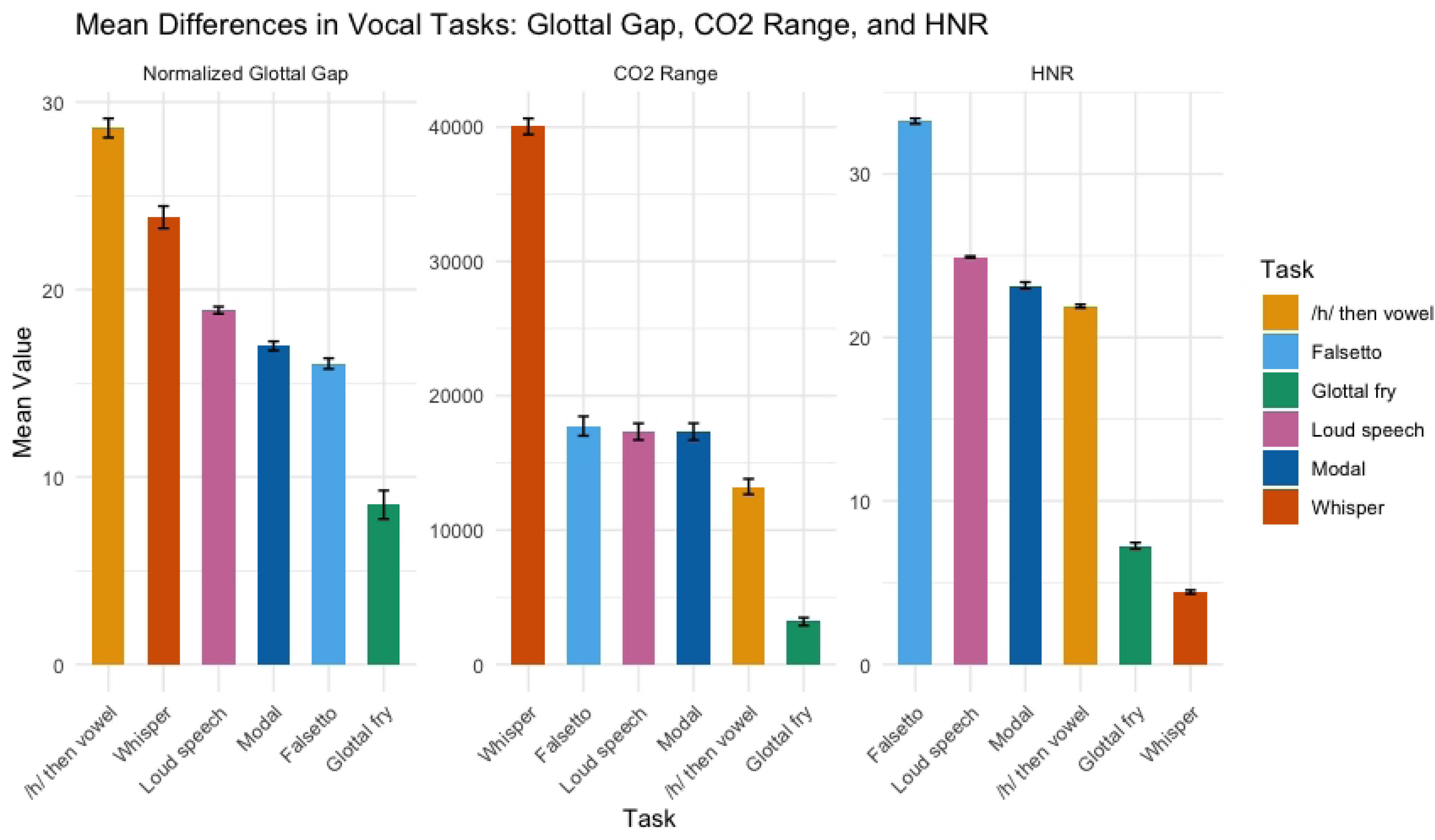
Mean differences in vocal tasks for Normalized Glottal Gap, CO_2_ Range, and HNR with standard error bars. Each bar represents the mean value for a specific vocal task, with tasks ordered from largest to smallest mean within each facet. Error bars indicate ±1 standard error. The x-axis lists the vocal tasks, and the y-axis shows the mean value for each variable. The CO_2_ Range label uses the correct subscript notation for CO_2_. Colors are consistent across facets, with each vocal task represented by the same color to facilitate comparison.

## Discussion

This study investigated how different phonation types influence aerosol generation, focusing on three key physiological factors: CO_2_ range, glottal gap, and harmonics-to-noise ratio (HNR). The results confirmed that whispering and loud speech produced the highest aerosol concentrations, while glottal fry generated the least. Smaller aerosol particles (0.1–1 µm) were more prevalent across all vocal tasks, highlighting their potential for airborne transmission.

However, whispering uniquely produced high emissions at both the smallest and largest particle sizes. Normalized glottal gap was the strongest predictor of aerosol generation, suggesting that increased vocal fold separation (such as in whispering and /h/ production) enhances airflow turbulence and particle formation. CO_2_ range was also a significant factor, suggesting that phonatory tasks that use a higher respiratory drive (such as loud voicing and whispering) drive more particle emissions. HNR played a lesser role, indicating that while periodicity contributes to particle production, it is not a primary mechanism.

Contrary to the assumption that quieter speech minimizes transmission risk, whispering generated the highest concentration of aerosol particles. This effect is likely due to the increased airflow turbulence through a partially opened glottis. Looking at **Figure 4**, whispering had the highest CO_2_ range across tasks and the second-largest normalized glottal gap. Therefore, the likely reason for the higher concentration of particles was the combination of both the higher airflow and the more open glottal posture. This finding is consistent with recent research from (11,12) showing higher aerosol concentration from whispered speech. An unexpected finding was that whispering produced a bimodal distribution of particles, with elevated emissions at both the smallest (0.1–1 µm) and largest (10–20 µm) particle size ranges. This dual-peak pattern differs from the typical gradual decline in concentration as particle size increases, observed for other phonation types. There are a few possible explanations for this generation of larger particles. First, the increased airflow turbulence through the glottis could have led to increases in vortex shedding and atomization, which could enhance the formation of both small and large droplets. Second, whispering involves a greater expiratory effort to maintain audibility without vocal fold vibration. This prolonged, turbulent airflow could promote the formation of larger particles, as exhaled moisture condenses and coalesces.

Also consistent with prior research, louder phonation resulted in greater aerosol concentration, likely driven by respiratory forces and higher amplitude of vocal fold vibration. Loud voicing requires higher subglottal pressure as well as stronger expiratory force, therefore requiring higher lung volume. Loud voicing particularly generated higher emissions at the smallest size range, with less pronounced differences across larger particle sizes. This finding is consistent with (8) showing higher particle concentrations for louder voicing.

Glottal fry produced the smallest concentration of droplets. This finding is consistent with the three physiological mechanisms studied: glottal fry had the smallest glottal gap, lowest CO_2_ range, and second-to-lowest HNR. This finding further reinforces the importance of these three variables, particularly airflow and glottal separation, in aerosol generation.

A critical aspect of this study was controlling for CO_2_ range, ensuring that variations in respiratory effort did not confound differences in aerosol concentration across phonation tasks. CO_2_ range served as a proxy for respiratory effort, with larger CO_2_ fluctuations indicating higher lung volume utilization and airflow velocity. By accounting for CO_2_ range in the statistical model, we isolated the effect of phonatory mechanisms (i.e., vocal fold vibration and glottal positioning) on aerosol generation. The high particle concentration across vocal tasks, despite controlling for CO_2_ range, underscores the role of airflow turbulence and glottal configuration in aerosol production. However, the finding that whispering, a voiceless phonation task, produced the highest concentration suggests that vocal fold vibration is not required for high aerosol output. Instead, airflow dynamics and glottal aperture size appear to be the primary drivers of aerosol generation, aligning with (10) who proposed that turbulent airflow, and not phonatory periodicity, enhances aerosolization.

These findings have important implications for airborne disease transmission. First, as found in (11,12), whispering is not necessarily a safer alternative for reducing aerosol emissions where airborne disease transmission is a concern. Rather, speaking with a clear, modal voice at a comfortable (and not loud) intensity level is likely to generate fewer particles. The high concentration of small particles (0.1–1 µm) in whispering and loud speech suggests that these vocal tasks may contribute significantly to airborne disease spread, as smaller particles remain suspended in air longer and travel further. The production of larger particles (10–20 µm) in whispering also raises concerns about fomite transmission, as these droplets can settle on surfaces.

While compelling, the results of this study had several limitations. First, the small sample size limits generalizability, and future studies should include larger, more diverse participant groups. Second, only sustained vowel production was used to attempt to isolate aerosol generation at the laryngeal level (and not from movement in the oral cavity). Future studies should examine aerosol production in naturalistic conversational speech, as individuals vary their speech intensity, airflow, and articulation dynamically. For example, projected speech in noisy environments (e.g., classrooms, restaurants) may increase aerosol emissions, while quiet, modal speech in one-on-one settings may minimize risk. Investigating these real-world speech variations would help refine public health guidelines for airborne disease mitigation in different social contexts. Finally, there is evidence that the APS is less efficient at measuring liquid particles 10 μm or higher (34), however it is not well understood if particles emitted from the respiratory tract are liquid or solid. We do know that they rapidly undergo the evaporation of some water and particle size shrinkage (35).

## Conclusion

This study highlights the significant role of airflow turbulence and glottal configuration in aerosol generation during phonation, with whispering and loud speech producing the highest particle concentrations and glottal fry generating the least. By controlling for CO_2_ range, we isolated the effects of phonatory mechanisms and found that vocal fold vibration alone is not the primary driver of aerosol production; instead, increased airflow turbulence and a more open glottal posture contribute to higher emissions. Notably, whispering produced a bimodal distribution of aerosol particles, with elevated concentrations at both the smallest and largest particle sizes, emphasizing its potential role in both airborne and fomite transmission. These findings reinforce the importance of considering speech-driven aerosolization in public health recommendations, particularly in high-density indoor environments where airborne disease transmission is a concern. Future research should explore conversational speech patterns and real-world speech variability to refine strategies for minimizing aerosol emissions in different communicative settings.

## Data Availability

All data and code files are available from the OSF database https://osf.io/uc6km/ DOI 10.17605/OSF.IO/UC6KM

https://osf.io/uc6km/10.17605/OSF.IO/UC6KM

## Acknowledgments (do not include funding here)

Thank you to Dr. Marie Jetté at the University of Colorado Anschutz Medical Campus Department of Otolaryngology for performing the laryngoscopy testing.

